# The causal relationship between gut microbiome composition and Alzheimer’s disease: A two-sample Mendelian randomization analysis

**DOI:** 10.1101/2025.08.20.25333769

**Authors:** Genevieve Monaghan, Charlie Hatcher, Emma L. Anderson, Kaitlin H. Wade

**Author notes:** Corresponding author Kaitlin H. Wade Oakfield House Oakfield Grove Bristol BS1 5LJ.

## Abstract

**Background:** The gut microbiome has been linked to multiple complex human traits and diseases, including Alzheimer’s disease (AD). However, it is unclear from existing studies whether these relationships are causal.

**Methods:** Two-sample bidirectional Mendelian randomisation (MR) was used to explore the causal relationship between the human gut microbiome with AD risk and two markers of cognitive function (fluid intelligence and reaction time). Sensitivity analyses explored validity of MR assumptions. We used data from the genome-wide association study (GWAS) meta-analysis of the gut microbiome (N=3890) in the Flemish Gut Flora Project (FGFP), Food-Chain Plus (FoCus) study and PopGen, two GWAS meta-analyses of late-onset, clinically diagnosed AD combined with proxy cases (N=455,258; clinically diagnosed cases only N=79,145), and GWASs of two cognitive function phenotypes in the UK Biobank: reaction time (N=459,523) and fluid intelligence (N=149,051).

**Results:** Initial results indicated that the abundance of an unclassified group of bacteria within the *Firmicutes* phylum decreased fluid intelligence and that presence vs. absence of bacteria within the *Dialister* genus increased the risk of AD (both clinical and proxy cases combined and clinical cases only). Five microbial traits had effect estimates that were directionally consistent across AD and cognitive phenotypes including those in the *Dialister, Parabacteroides and Ruminococcus* genera, the *Firmicutes* phylum and the *Porphyromonadaceae* family. Sensitivity analyses indicated that our results were likely biased either by horizontal pleiotropy, genetic confounding or reverse causation and, thus, unlikely to reflect a causal relationship, further highlighting the importance of conducting such sensitivity analyses and caution in causal interpretation.

**Conclusions:** Whilst our analyses initially provided evidence that features of the gut microbiome may influence the risk of AD and cognition, further sensitivity analyses indicated that these results were likely not reflective of causality.

## Background

Dementia is the most common cause of death in the UK and prevalence is rising, with 152.8 million people estimated to be living with dementia by 2050 (1). Alzheimer’s disease (AD) is the most common cause of dementia and is associated with memory loss and reduced cognitive function, including slower reaction times and lower fluid intelligence (1–3). AD risk increases with age, but it is also highly heritable, with 75 common single nucleotide polymorphisms (SNPs) identified thus far in genetic studies (2, 3).

The gut microbiome has been linked to human health outcomes across multiple complex traits and diseases, including AD (4, 5). Multiple observational studies have linked AD to disturbances in the gut microbiome, although the specific alterations to gut microbiome abundance varies between studies (6–11). With these inconsistencies in the current evidence, it is unclear whether the relationship between the gut microbiome and AD is causal or biased (e.g. by confounding). It is also plausible that the inconsistencies observed in relationships between the gut microbiome and AD are explained by reverse causality or a bidirectional relationship. For example, one large observational study found lower microbial diversity in cases of mild cognitive impairment (a precursor to dementia) compared to the healthy controls, coupled with lower abundance of bacteria within the *Faecalibacterium* genus, *Ruminococcaceae* family, and *Alistipes* genus and a higher abundance of bacteria within the *Proteobacteria* phylum and *Gammaproteobacteria* class (7). Whilst another study found similarly low microbial diversity in cases of mild cognitive impairment compared to controls, the only differences in specific microbial traits were a lower abundance of bacteria in the *Parabacteroides* genus and a higher abundance of bacteria in the *Dialister* genus (10). The effect of the gut microbiome on AD and the neuroinflammatory mechanisms through which this effect may occur was explored further though a recent review which suggested that changes to the gut microbiome, such as reduced abundance of bacteria of the *Bifidobacterium* genus *Firmicutes* phylum, leads to increased permeability of the lining of the gut which increases neuroinflammation via the gut-brain axis (11).

Mendelian randomization (MR) is an instrumental variable method that uses genetic variants as instruments for the exposure, to investigate the causal effect of the exposure on an outcome (12, 13). Previous MR studies have linked AD to other diseases such as epilepsy, and linked neuroinflammation to AD, among other findings (3, 14, 15). More recently, MR has also been applied to understand the causal role of the gut microbiome in AD risk (16–19). However, the findings from these studies are very likely biased due to the use of lenient p-value thresholds for instrument selection, and little investigation of horizontal pleiotropy. Therefore, results from existing MR studies of the gut microbiome should be interpreted with caution (5).

In this paper, we aimed to appraise the current application of MR in the gut microbiome field within the context of exploring the causal relationship between features of the human gut microbiome and AD risk and cognitive function phenotypes, including fluid intelligence and reaction time. We undertook two-sample bidirectional MR analyses using summary-data from publicly available genome-wide association studies (GWASs) of all phenotypes, selecting instruments for our exposures using robust p-value thresholds and undertaking a comprehensive series of sensitivity analyses to interrogate the validity of the core MR assumptions.

## Methods

### Study design

We conducted two-sample bidirectional MR analyses using summary-level data from some of the largest GWAS of the gut microbiome, AD, and two cognitive function phenotypes (reaction time and fluid intelligence). For clarity, analyses exploring the causal role of the gut microbiome on AD risk and related cognitive phenotypes are referred to as analyses in the “forwards direction”, and analyses exploring the effect of AD liability and related cognitive phenotypes on the gut microbiome are referred to as analyses in the “reverse direction”.

### GWAS summary data and instrument selection

We used data from one of the largest GWAS meta-analyses of the gut microbiome (N=3,890), which included data from three independent non-overlapping GWASs (the Flemish Gut Flora Project [FGFP; N=2,223], the Food-Chain Plus study [FoCus; N=950], and the PopGen study [N=717]) (20). Full details on the cohorts, sampling, and analyses have been previously described (20–22). In summary, the hypervariable regions of the 16S rRNA gene (V4 hypervariable region for FGFP and V1-V2 hypervariable regions for FoCus and PopGen) and DNA extracted from the participants’ frozen faecal samples were amplified and sequenced (20–22). To provide bacterial taxonomic classifications for the microbiota, for each sample the resultant sequences were analysed using the DADA2 pipeline (20). In cases where the microbiota could not be confidently classified at the genus level, they were organised into arbitrary unclassified groups within higher classification levels (20). Informed consent and appropriate ethical approval were obtained for each study (20–22). A genome-wide p-value threshold (P < 2.5e-8), accounting for the number of independent microbial traits analysed in the GWAS, was used to define the microbiome instruments from the genome-wide meta-analysis for the forwards direction (i.e. of the gut microbiome on AD and related phenotypes) (4). Using these criteria, 13 microbial traits had one associated SNP that reached the genome-wide p-value threshold in the microbiome GWAS meta-analysis. In addition, one SNP that was associated with bacteria within the *Bifidobacterium* genus was also included in analyses which, although not reaching the genome-wide p-value threshold in the meta-analysis, has been consistently reported across independent studies as being associated with *Bifidobacterium* bacteria (20). Therefore, a total of 14 SNPs were used as genetic instruments in our forwards MR analyses, with one SNP being associated with each microbial trait (20).

For AD, we used GWAS summary statistics from Jansen et al. (2019), which is a large publicly available GWAS of AD. This GWAS was selected as it is comprised of three phases: phase 1 (clinically diagnosed late-onset AD cases [N=24,087 cases/55,058 controls]), phase 2 (“proxy” AD cases [N=47,793 cases/328,320 controls]), and phase 3 (meta-analysis of clinical and “proxy” AD cases of European ancestry from phase 1 and phase 2 [N=71,880 cases/383,378 controls]) (2). Clinically diagnosed cases were comprised of patients from the AD Sequencing Project, the International Genomics of Alzheimer’s Project (IGAP), and the AD working group of the Psychiatric Genomics Consortium (PGC) (2). Patients were regarded as “proxy” cases if they responded affirmatively to the question of whether their father or mother had ever suffered from dementia/AD in UK Biobank (2). We used the GWAS summary statistics from phases 1 and 3 in our analyses to enable a comparison of results when the less reliable “proxy-cases” were included, whilst boosting sample size.

We additionally used two continuous cognitive function measurements from the UK Biobank: reaction time (N=459,523) and fluid intelligence (N=149,051) (see **Supplementary Information 1**) (3). Summary statistics for both cognitive phenotypes were obtained through the IEU OpenGWAS pipeline (23).

For the analysis in the reverse direction (i.e. AD liability and related cognitive phenotypes on the gut microbiome), SNPs that met the genome-wide significance p-value threshold (P < 5) were selected as instruments from each respective GWAS summary-level data. A total of 176 fluid intelligence SNPs, 76 reaction time SNPs, 19 AD meta-analysis SNPs, and 18 clinical AD SNPs were used as genetic instruments in our reverse MR analyses (20).

### MR analyses

For the primary MR analysis, bidirectional two-sample MR was performed to investigate the causal relationship between 14 microbial traits with AD risk, fluid intelligence and reaction time, using the TwoSampleMR package (version 0.5.7 in R version 4.3.0). For the forwards direction, summary-level statistics (i.e. SNP name, effect allele, other allele, effect allele frequency, beta coefficient, standard error, p-value, and sample size) were extracted for each of the 14 microbial trait-associated SNPs from the GWAS of the microbiome, both AD GWAS, and the fluid intelligence and reaction time GWAS. No SNPs were excluded for being palindromic with ambiguous allele frequencies in the forwards direction. For the reverse direction, the same summary-level statistics were extracted for all SNPs associated with both AD traits, fluid intelligence and reaction time from all GWAS, including the GWAS meta-analysis of the 14 gut microbial traits used in the forwards direction. In both forwards and reverse MR analyses, the exposure and outcome datasets were harmonised so that the effect of each SNP on both the exposure and outcome was relative to the same effect allele using information on the effect allele and effect allele frequencies.

In the forwards direction, as there was only one associated SNP per microbial trait, Wald ratios were used for the main analysis assessing the causal role of the microbiome on the AD and cognitive phenotypes. For reverse direction, the inverse variance weighted (IVW) method was used as the main analysis as there were multiple SNPs associated with AD and cognitive phenotypes. Assuming that the AD and cognitive phenotypes were highly correlated, p-values for MR analyses were corrected for multiple testing using the Bonferroni method, adjusting for the number of microbial traits (P = 0.05/14 = 0.0036) in each direction, separately.

In addition to looking at each exposure-outcome relationship individually, in the forwards direction we compared patterns of results across AD outcomes to investigate whether all effect estimates for any microbial trait were consistently beneficial or detrimental to AD risk – we refer to this in the results as “biologically directionally consistent”. Whilst for both AD traits and reaction time, higher values reflect poorer outcomes and greater risk of AD, higher effect estimates for fluid intelligence reflected better cognitive function. Thus, to aid in interpretation, these values were inversed to be biologically directionally consistent with the other three cognitive phenotypes so that greater effect estimates reflect poorer fluid intelligence. MR estimates in the forwards direction are interpreted as the average difference in odds of AD risk, or the average standard deviation (SD) change in cognitive phenotypes, either per SD higher abundance of continuous microbial traits, or per an approximate doubling of genetic liability to each binary presence vs. absence microbial trait. In the reverse direction, MR estimates can be interpreted as the average SD change in each continuous microbial trait, or the average difference in odds for presence vs. absence in each binary microbial trait, either per SD higher fluid intelligence score or reaction time, or per approximate doubling of genetic liability to AD. This study and all methods have been conducted in line with the Strengthening the Reporting of Observational Studies in Epidemiology MR (STROBE-MR) reporting guidelines for MR studies and a full checklist is provided in the **supplement** (24).

### Sensitivity analyses

When assessing whether MR estimates likely represent a causal effect of an exposure on an outcome, three core assumptions must be met. Firstly, the genetic instruments selected as instruments for the exposure must be strongly associated with the exposure (IV1) (25). Secondly, there should be no confounding between the genetic instruments and the outcome (IV2) (25). And thirdly, the genetic instruments should have no effect on the outcome, other than via the exposure (i.e. no horizontal pleiotropy) (IV3) (25). We explored the validity of these assumptions in our analyses and the robustness of our findings by performing an array of sensitivity analyses, including an exploration of horizontal pleiotropy and colocalisation. The description of these sensitivity analyses is provided below, separated into those used in the forwards and reverse directions.

### Forwards direction

Firstly, genetic colocalisation was conducted to assess whether any observed effect between the gut microbiome and AD risk or cognitive function was driven by a shared causal variant; a necessary (but not sufficient) criterion for causality. The ‘coloc’ R package was used with default parameters to investigate colocalisation for each microbial trait for which there was evidence of a causal effect on AD risk and/or cognitive phenotypes in the forwards MR, with cognitive function measure fluid intelligence and microbial abundance trait Firmicutes specified as continuous variables (“quant”), and microbial presence vs. absence trait Dialister and both AD traits specified as binary (“cc”). Five posterior probabilities (H0-H4) were generated using Bayes factor computation: (H0) neither trait has a genetic association in the region; (H1) only the gut microbiome has a genetic association in the region; (H2) only AD or the cognitive function measures has a genetic association in the region; (H3) both traits are associated but have different causal variants; and (H4) both traits are associated and have the same causal variant. We used a posterior probability threshold ofl1≥l10.80 to indicate evidence of a shared common causal variant between each microbial trait and AD-related cognitive phenotypes. However, due to the likely low power in these analyses and given sample size of the gut microbiome GWAS data (i.e. where we only had genome-wide data available for the FGFP cohort), we compared H1 vs. H2 and H4 vs. H3 as forms of suggestive evidence for causality. For example, if H4 was greater than H3, there was greater chance of colocalisation (H4) vs. a different causal variant (H3), and if H1 was greater than H2, there was greater chance that the genetic variant was predominantly associated with the exposure (H1; which is assumed given the nature of instrument selection) rather than being more associated with the outcome (H2; which would bias MR estimates given that the SNP may be a more relevant instrument for the outcome than the exposure). To complement the colocalisation analyses, we also performed the Steiger test to check that the genetic instruments explained more variance in the microbial traits, than they did in AD risk or the cognitive phenotypes.

Secondly, given that there were only singular SNPs associated with each microbial trait, we were unable to perform pleiotropy-robust methods that require multiple genetic instruments, such as MR-Egger (26). Therefore, for the forwards direction, we manually explored the likelihood of horizontal pleiotropy by searching the IEU OpenGWAS for SNPs associated with each microbial trait for which there was evidence of a causal effect on AD risk and/or cognitive phenotypes, to identify whether these SNPs were associated with any trait that could be directly or indirectly related to AD and related phenotypes (23). To identify SNPs strongly associated with phenotypes in the IEU OpenGWAS, a lenient p-value (P <l11e-05) was used for these exploratory sensitivity analyses.

Thirdly, to allow the use of more formal pleiotropy-robust methods in the forwards direction, a lenient p-value threshold (P <l11e-05) was used to select genetic instruments for each microbial trait for which there was evidence of a causal effect on AD risk and/or cognitive phenotypes when using the stricter p-value threshold. After selecting SNPs based on the lenient p-value threshold, we then restricted these associated SNPs to those that had directionally consistent effect estimates across each of the three cohorts included in the GWAS of the gut microbiome (4). This increase in the number of instruments then allowed us to perform several sensitivity analyses investigating unbalanced horizontal pleiotropy including the MR-Egger, weighted median, and weighted mode methods (see **Supplementary Information 2**) (26–28).

### Reverse direction

Given the existence of multiple SNPs associated with both AD liability and cognitive phenotypes, we estimated the level of heterogeneity of MR estimates across all SNPs using the Cochran’s Q statistic and the likelihood of unbalanced horizontal pleiotropy using the same pleiotropy-robust methods and sensitivity analyses as above.

## Results

### Instrumentation

Full information on the SNPs associated with the gut microbiome can be found in **Table 1**. The F statistics (range = 0.082 – 176.3) suggest that the likelihood of weak instrument bias was high in *F. Sutterellaceae* (presence/absence [P/A]), *G. Coprococcus* (P/A), *G. Ruminococcus* (P/A), and *G. u. P. Firmicutes* (P/A). In the forwards direction, all 14 of the microbiome-related SNPs were available for analysis with the AD and cognitive phenotypes.

**Table 1.** Host genetic variants associated with features of the human gut microbiome. The 14 human genetic variants associated features of the human gut microbiome identified from the largest genome-wide association study published by Hughes et al. 2020. For each microbial trait, the small nucleotide polymorphism (SNP), effect allele (EA), other allele (OA), EA frequency (EAF) are given. The betas and corresponding standard errors (SE) represent the odds ratio for AD risk per standard deviation unit change for continuous microbial traits (labelled as “AB”) or per approximate doubling of the genetic liability to presence of each binary microbial trait (labelled as “P/A”). The variance explained in the exposure by the associated SNP is represented by R and the F-statistic gives an indicator of the strength of each instrumental variable. The letters in the microbial trait name represent the taxon classification which each microbial trait was observed at with “P”, “O”, “F”, and “G” representing “phylum”, “order”, “family”, and “genus”, respectively. Any microbial trait which could not be confidently classified at the genus level were labelled “unclassified” (“u”) and then organised within a higher classification rank.

| SNP | Microbial Trait | OA | EA | EAF | Beta (SE) | p-value | N | R<sup>2</sup> | F |
| --- | --- | --- | --- | --- | --- | --- | --- | --- | --- |
| rs116865000 | *C. Gammaproteobacteria* (AB) | G | A | 0.03 | 0.56 (0.10) | 8.41E-09 | 1626 | 0.02 | 33.38 |
| rs4988325 | *G. Bifidobacterium* (AB) | G | A | 0.65 | -0.13 (0.03) | 1.34E-06 | 1975 | 0.01 | 18.76 |
| rs55808472 | *G. Butyricoccus* (AB) | G | A | 0.07 | 0.26 (0.04) | 5.54E-10 | 2223 | 0.02 | 39.26 |
| rs13207588 | *G. Parabacteroides* (AB) | G | A | 0.23 | -0.18 (0.03) | 6.80E-09 | 2223 | 0.01 | 33.68 |
| rs35980751 | *G. u. F. Porphyromonadaceae* (AB) | G | T | 0.26 | 0.20 (0.03) | 5.07E-09 | 1420 | 0.02 | 33.19 |
| rs11788336 | *G. u. P. Firmicutes* (AB) | T | C | 0.27 | -0.14 (0.03) | 1.66E-08 | 1904 | 0.02 | 32.68 |
| rs4494297 | *F. Sutterellaceae* (P/A) | G | T | 0.01 | 0.14 (0.31) | 6.80E-10 | 2223 | 6.86E-05 | 0.15 |
| rs561177583 | *G. Coprococcus* (P/A) | G | A | 0.01 | 0.16 (0.28) | 1.10E-10 | 2223 | 9.34E-05 | 0.21 |
| rs7118902 | *G. Dialister* (P/A) | G | A | 0.31 | 0.73 (0.05) | 4.14E-09 | 2223 | 0.03 | 77.24 |
| rs150018970 | *G. Ruminococcus* (P/A) | G | A | 0.01 | 0.11 (0.29) | 6.68E-14 | 2223 | 3.71E-05 | 0.08 |
| rs6733298 | *G. u. F. Erysipelotrichaceae* (P/A) | A | G | 0.89 | 1.64 (0.09) | 6.91E-09 | 2223 | 0.07 | 176.34 |
| rs116135844 | *G. u. O. Bacteroidales* (P/A) | G | A | 0.04 | 2.11 (0.13) | 2.32E-08 | 2223 | 0.05 | 123.57 |
| rs34656657 | *G. u. P. Firmicutes* (P/A) | G | A | 0.02 | 0.29 (0.22) | 1.82E-08 | 2223 | 5.65E-04 | 1.26 |
| rs117338748 | *G. Veillonella* (P/A) | G | A | 0.02 | 2.89 (0.19) | 2.42E-08 | 2223 | 0.05 | 104.88 |

### MR analyses

In the forwards direction, there was evidence that a higher abundance of an unclassified group of bacteria within the *Firmicutes* phylum (*G. u. P. Firmicutes* abundance [AB]) decreased fluid intelligence (SD change in fluid intelligence per one SD higher abundance [Wald ratio] = 0.80, standard error [SE] = 0.07, p-value = 0.0029). There was also evidence that the presence vs. absence of bacteria in the *Dialister* genus (*G. Dialister* [P/A]) increased the risk of AD both with clinical and proxy cases combined (OR with an approximate doubling of the genetic liability to presence vs. absence = 1.01, SE = 0.003, corrected p-value = 0.004) and with clinical cases only (OR = 1.07, SE = 0.02, corrected p-value = 0.01, see **Figure 1** and **Table 2**).

**Figure 1.**
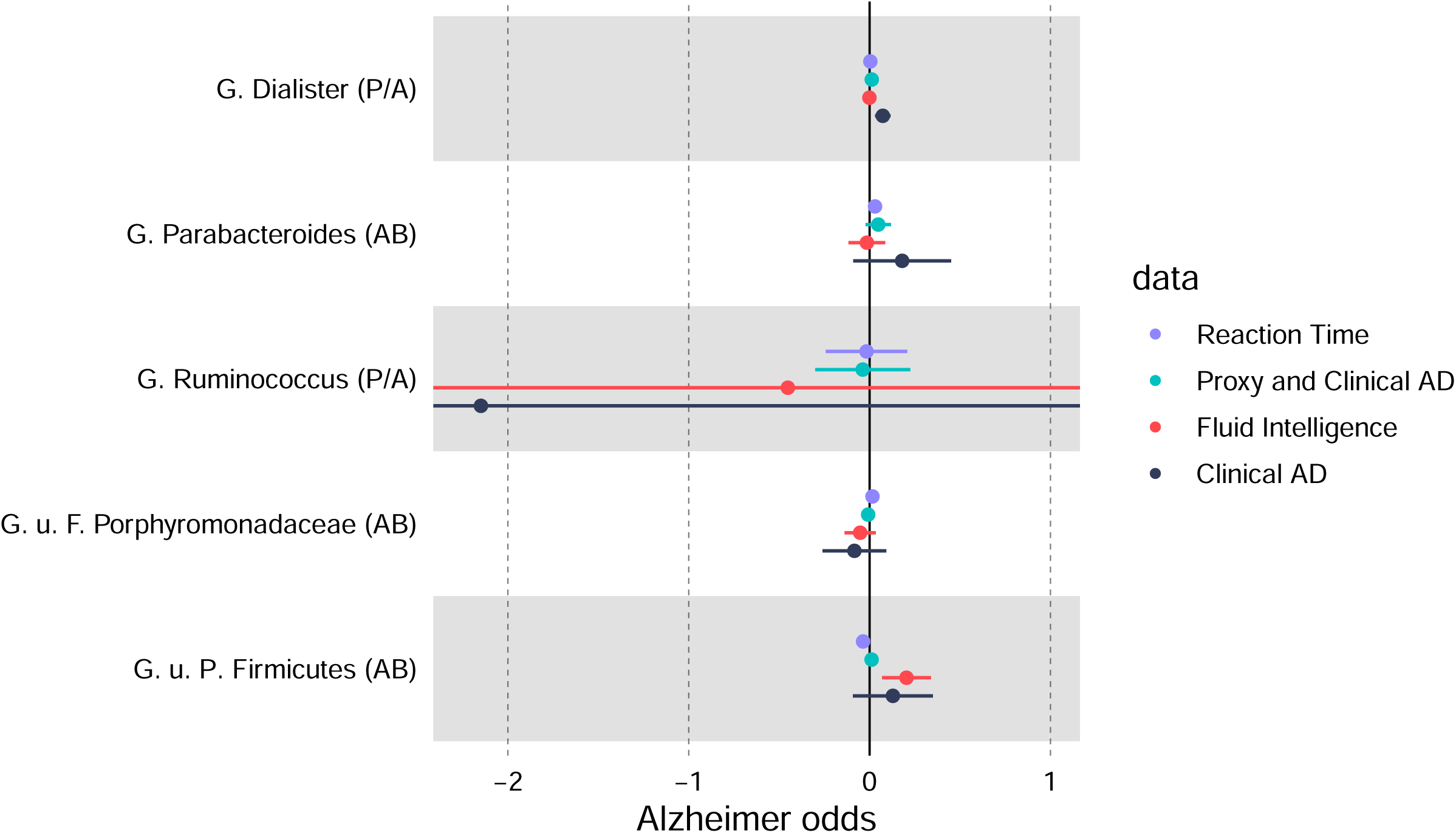
MR estimates of the effect of select microbial trait on AD and cognitive intelligence phenotypes. The letters in the microbial trait name represent the taxon classification which each microbial trait was observed at with “P”, “F”, and “G” representing “phylum”, “family”, and “genus”, respectively. All microbial traits which could not be confidently classified at the genus level were labelled “unclassified” (“u”). MR estimates represent the odds ratio for AD risk and 95% confidence intervals per standard deviation unit change for continuous microbial traits (labelled as “AB”) or per approximate doubling of the genetic liability to presence of each binary microbial trait (labelled as “P/A”). The estimate for clinical AD for *G. Ruminococcus* (P/A) had a substantially larger estimate than the others with such wide confidence intervals that it was not possible to show on the figure and still maintain a sufficient level of detail for the rest of the estimates.

**Table 2.**
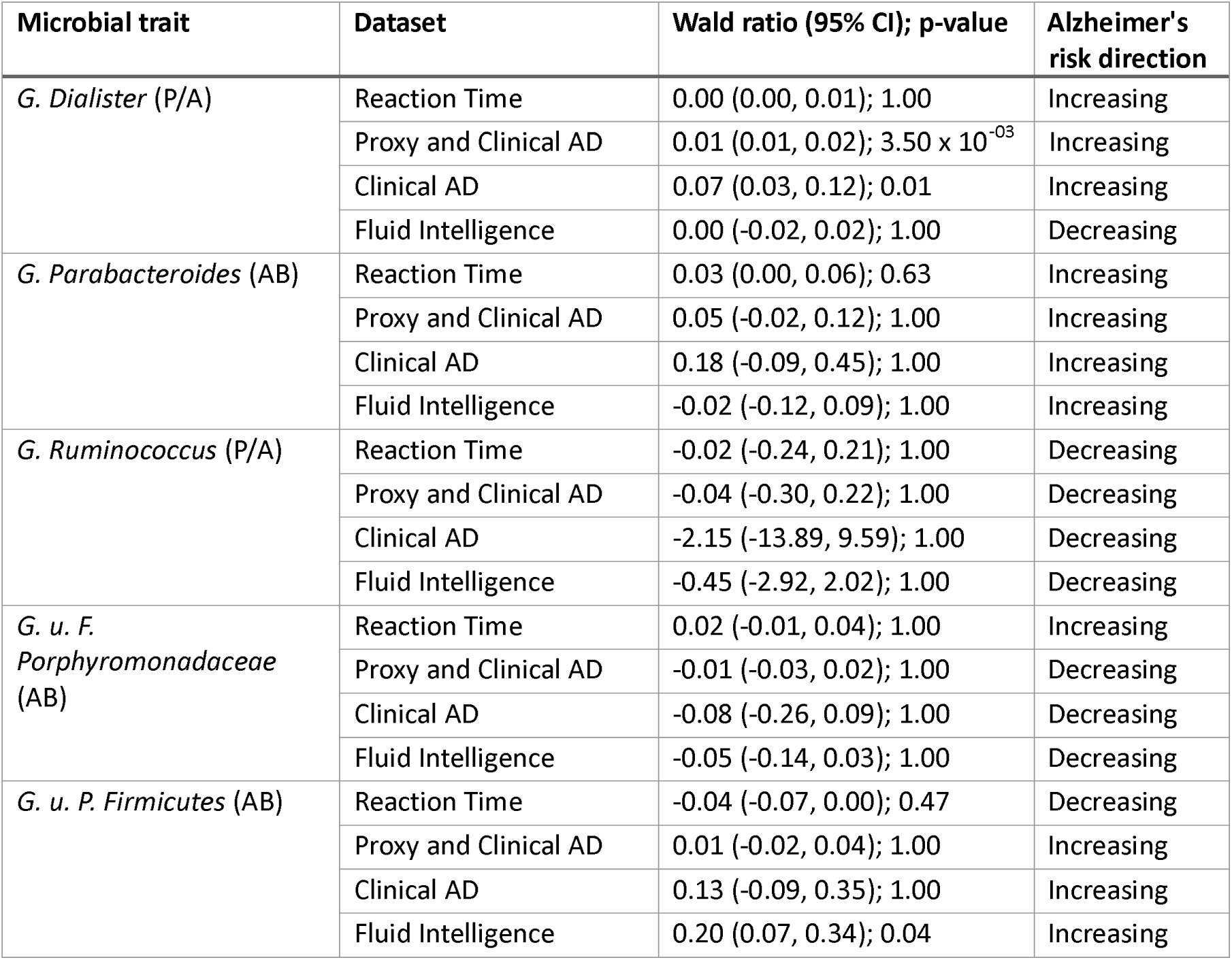
Two-sample MR estimates of the effect of microbial traits on AD risk. The letters in the microbial trait name represent the taxon classification which each microbial trait was observed at with “P”, “F”, and “G” representing “phylum”, “family”, and “genus”, respectively. Any microbial trait which could not be confidently classified at the genus level were labelled “unclassified” (“u”) and then organised within a higher classification rank. The MR effect estimates represent the odds ratio for AD risk and 95% confidence intervals per standard deviation unit change for continuous microbial traits (labelled as “AB”) or per approximate doubling of the genetic liability to presence of each binary microbial trait (labelled as “P/A”).

| Microbial trait | Dataset | Wald ratio (95% CI); p-value | Alzheimer's risk direction |
| --- | --- | --- | --- |
| *G. Dialister* (P/A) | Reaction Time | 0.00 (0.00, 0.01); 1.00 | Increasing |
| | Proxy and Clinical AD | 0.01 (0.01, 0.02); $3.50 \times 10^{-03}$ | Increasing |
|  | Clinical AD | 0.07 (0.03, 0.12); 0.01 | Increasing |
|  | Fluid Intelligence | 0.00 (-0.02, 0.02); 1.00 | Decreasing |
| *G. Parabacteroides* (AB) | Reaction Time | 0.03 (0.00, 0.06); 0.63 | Increasing |
|  | Proxy and Clinical AD | 0.05 (-0.02, 0.12); 1.00 | Increasing |
|  | Clinical AD | 0.18 (-0.09, 0.45); 1.00 | Increasing |
|  | Fluid Intelligence | -0.02 (-0.12, 0.09); 1.00 | Increasing |
| *G. Ruminococcus* (P/A) | Reaction Time | -0.02 (-0.24, 0.21); 1.00 | Decreasing |
|  | Proxy and Clinical AD | -0.04 (-0.30, 0.22); 1.00 | Decreasing |
|  | Clinical AD | -2.15 (-13.89, 9.59); 1.00 | Decreasing |
|  | Fluid Intelligence | -0.45 (-2.92, 2.02); 1.00 | Decreasing |
| *G. u. F. Porphyromonadaceae* (AB) | Reaction Time | 0.02 (-0.01, 0.04); 1.00 | Increasing |
|  | Proxy and Clinical AD | -0.01 (-0.03, 0.02); 1.00 | Decreasing |
|  | Clinical AD | -0.08 (-0.26, 0.09); 1.00 | Decreasing |
|  | Fluid Intelligence | -0.05 (-0.14, 0.03); 1.00 | Decreasing |
| *G. u. P. Firmicutes* (AB) | Reaction Time | -0.04 (-0.07, 0.00); 0.47 | Decreasing |
|  | Proxy and Clinical AD | 0.01 (-0.02, 0.04); 1.00 | Increasing |
|  | Clinical AD | 0.13 (-0.09, 0.35); 1.00 | Increasing |
|  | Fluid Intelligence | 0.20 (0.07, 0.34); 0.04 | Increasing |

There was little evidence for an effect of other microbial traits on any of the AD or related phenotypes. However, five microbial traits did have effect estimates that were biologically directionally consistent across both AD phenotypes and one or both cognitive intelligence phenotypes. Specifically, the abundance of *G. Parabacteroides* and *G. u. P. Firmicutes* as well as the presence vs. absence of *G. Dialister* consistently increased AD risk and lowered cognitive function, whilst the presence of *G. Ruminococcus* and the abundance of *G. u. F. Porphyromonadaceae* consistently decreased AD risk and increased cognitive function (see **Figure 1**). *G. Ruminococcus* (P/A) was the only phenotype that was biologically and directionally consistent across all four phenotypes. Further information on effect estimates for all 14 microbial traits for all four outcomes can be found in **Supplementary Table 1**.

Reverse MR analysis found little evidence for an effect of AD or the cognitive phenotypes on the gut microbiome after adjusting for multiple testing, however the strongest associations observed were between fluid intelligence and *F. Enterococcaceae* (AB) (beta = 0.406, P = 0.001), and Reaction Time and *G. Desulfovibrio* (P/A) (beta = 2.028094, P = <0.001).

### Sensitivity Analyses Forwards direction

Colocalisation analyses suggested that the effect of the gut microbiome on AD risk was unlikely to be causal and instead due to different causal variants in the same genomic region or linkage disequilibrium (LD) (see **Supplementary Table 2** and **Figure 2**). Specifically, colocalisation analyses showed that there were likely different causal variants in the region surrounding the top SNP associated with *G. Dialister* (P/A) (rs7118902) for AD risk (posterior probability [P] for H3 = 0.996 for both AD GWAS). Furthermore, H2 was greater than H1 for both phenotypes, suggesting that the SNP used as an instrument for *G. Dialister* (P/A) in the forwards direction was more associated with the AD phenotypes. For *G. u. P. Firmicutes* (AB), although the posterior probability for H4 (P = 0.48) at the most related SNP (rs118011482) was greater than H3 (P = 0.17) and H1 (P = 0.34) was greater than H2 (P = 0.0001), the regional plots showed a high level of LD within the region, suggesting that the original causal effect may be due to genetic confounding (see **Figure 2c**).

**Figure 2.**
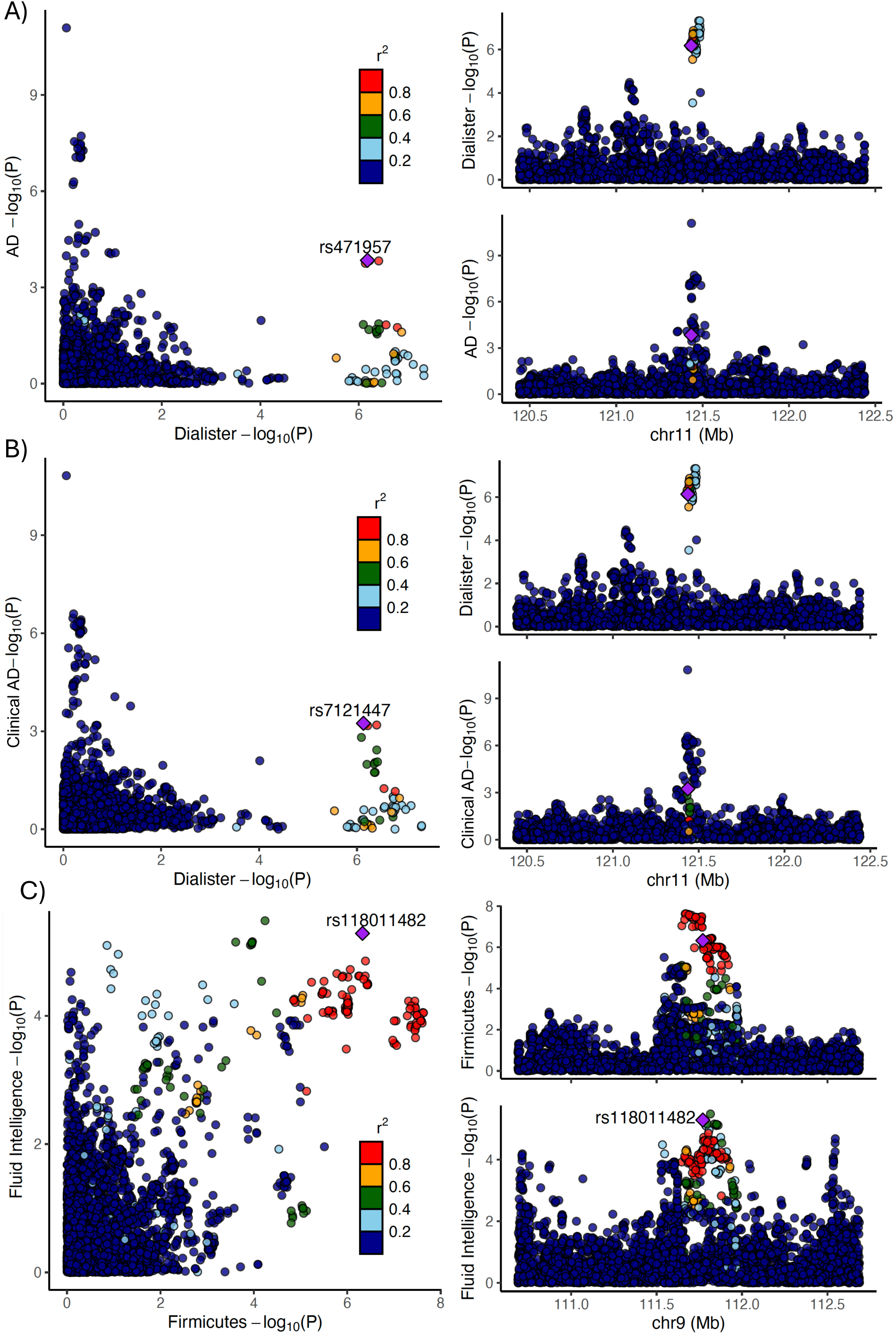
Colocation results. Colocalisation results for A) *G. Dialister* (P/A) microbial trait with Proxy and Clinical meta-analysed AD; B) *G. Dialister* (P/A) microbial trait with Clinical AD; and C) *G. u. P. Firmicutes* (AB) with fluid intelligence. The regional association plots show − log10(P-value) where each lead SNP is represented by a purple diamond in relation to AD or fluid intelligence. The letters in the microbial trait name represent the taxon classification which each microbial trait was observed at with “P” and “G” representing “phylum” and “genus”, respectively. Any microbial trait which could not be confidently classified at the genus level were labelled “unclassified” (“u”) and then organised within a higher classification rank.

[Steiger filtering results indicated that for all SNPs but one, the causal direction was from the gut microbiome to AD, and the SNP explained more variance in the microbial trait than the outcome (see **Supplementary Table 3**). For all three SNPs for which there was evidence of a causal association between the gut microbiome and AD in the forwards MR, the Steiger filtering results indicated that the direction of this association was from the microbial trait to AD risk. *G. Ruminococcus* (P/A) was the only SNP for which there was suggestive evidence that the SNP explained more variance in AD risk than in the microbial trait (i.e. indicative of causality in the reverse direction), however P = 0.88.

The IEU OpenGWAS identified 58 traits associated with the SNP used as the instrument for *G. Dialister* (P/A) (rs7118902) including 3mm weak meridian (right) (P = 3.10e-10), 6mm weak meridian (right) (P = 8.70e-10) (23). Similarly, 154 traits were associated with the SNP related to *G. u. P. Firmicutes* (AB) (rs11788336) in IEU OpenGWAS, including fluid intelligence (P = 0.00047) and anthropometric measures such as weight (P = 2.46e-11) and body fat percentage (P = 0.00082), all of which may be indicative of pleiotropy (i.e. independent pathways between the SNPs associated with both microbial traits and AD risk) or reverse causality (23).

When a more lenient p-value threshold (P <l11e-05) was used to select genetic instruments for the five microbial traits which had a biologically directionally effect on AD risk, 19 SNPs across the five microbial traits were included (compared to 5 with the more stringent p-value threshold) (see **Supplementary Table 4**). The only result with Bonferroni P < 0.05 was a negative relationship between AD in the meta-analysis of proxy and clinical cases and *G. Dialister* (P/A) (rs7118902).

### Reverse direction

The MR Egger, IVW, weighted median, and weighted mode analyses all showed that there was limited evidence of horizontal pleiotropy in the reverse MR; intercepts were all close to zero (see **Supplementary Table 5**).

## Discussion

We investigated the relationship between 14 microbial traits and AD risk (both clinically diagnosed and clinically diagnosed combined with proxy indicators) and two measures of cognitive function (fluid intelligence and reaction time). Our initial analyses provided evidence that a higher abundance of bacteria within the *Firmicutes* phylum decreased fluid intelligence and that presence of bacteria within the *Dialister* genus increased the risk of AD when defined either by a clinical diagnosis or with a combination of clinical diagnoses and by proxy. Importantly, however, after undertaking a series of sensitivity analyses that interrogated possible violations of MR assumptions, it is clear that these initial findings are unlikely to reflect causality, and more likely be a function of reverse causality or genetic confounding.

The colocalisation analyses indicated that the results may be explained by genetic confounding (i.e. where the “microbiome-related SNP” is highly correlated with an “AD-related SNP”, biasing the MR estimate). Consistent with this, our manual exploration of horizontal pleiotropy suggested that SNPs associated with these microbial traits were also highly related to a collection of phenotypes that could additionally be related to AD – either directly or indirectly via other phenotypic pathways. Lastly, and particularly in the case of the *Dialister* microbial trait, the colocalisation showed that the SNP related to this trait in the GWAS meta-analysis was much more strongly related to, and explained more variation in, AD risk, implying that the MR effect estimate was very likely explained by reverse causality (i.e. where the “microbiome-related SNP” impacts the microbiome via AD instead of vice versa). Supportive of this, it is worth noting that the SNP associated with the *Dialister* microbial trait is near a known AD-related gene (*SORL1*) (29). That said, this genomic colocalisation does not imply a definitive biological relationship with that gene and reverse MR analyses and Steiger filtering did not provide strong evidence for a causal effect of AD and cognitive phenotypes on these microbial traits (though this may have been limited by the statistical power within those analyses, given the relatively small sample size of the microbiome GWAS). However, the observed effects may indeed be explained by the complexity of genetic architecture shared across these phenotypes rather than a causal effect in either direction.

Overall, this work highlights the importance of using methods like colocalisation and a more comprehensive set of sensitivity analyses, in questioning the validity of obtained results coupled with a greater understanding of the genetic variants being used in MR analyses of the gut microbiome, particularly when the number of available genetic variants is small. What is required in all future applications of MR analyses that attempt to understand the causal role played by the gut microbiome (or indeed any exposure) in health and disease is to treat the initial results with caution. Related to this, several previous MR studies investigating the causal role of the gut microbiome on risk of AD have presented inconsistent effect estimates despite using similar techniques and, in some cases, the same data (16–19). The lack of consistency across multiple MR analyses of the gut microbiome and AD reaffirms the need for greater caution in applying these analyses and interpreting effect estimates carefully.

In our exploration of consistency in effect estimates across AD and cognitive phenotypes with each microbial trait, we did observe directional (and, thus, potentially biologically relevant) consistency across only one of the microbial traits analysed (the *Ruminococcus* genus). However, there was a lack of strong statistical evidence for a causal relationship in these analyses, potentially due to a lack of statistical power. Therefore, it may be worth investigating the biologically and directionally consistent results further, appropriately applying MR analyses using larger GWAS of the microbiome and triangulating this evidence across various study designs with orthogonal sources of bias (30). However, it is possible that these consistent directions of effect could be also explained by the aforementioned complexities and challenges.

Our study demonstrates the utility of appropriately applying bidirectional two-sample MR in the context of understanding the causal role of the gut microbiome on AD. Whilst many other MR studies centring around the gut microbiome tend to use lenient p-value thresholds for instrument selection, our analysis used genetic variants that met the genome-wide p-value threshold (P < 2.5) in the GWAS meta-analysis of the gut microbiome. Even though this is best practice in MR analyses (as it ensures that SNPs used are robustly associated with each microbial trait and, thus, adheres to the first MR assumption, IV1), our work demonstrates that violations of the core MR assumptions are still possible when using such complex phenotypes even when using the most appropriate genetic variants. Thus, analyses using lenient p-value thresholds (e.g. the most commonly used threshold of <1 among MR papers focusing on the gut microbiome) should be considered unreliable. Though a lenient p-value threshold may increase the variance explained in microbial traits, it yields instruments that violate at least the IV1 assumption (as variants are not robustly associated with the microbial traits) and also increases risk of violation of IV3. This threatens our ability to make causal inferences and, thus, undertaking the types of sensitivity analyses presented here are recommended.

A recent systematic review and meta-analysis of AD and gut microbiome found that some of these differences between studies were the result of different clinical stages of AD and the setting of the study; although decreased microbial diversity, increased abundance of bacteria of the *Proteobacteria* phylum and *Bifidobacterium* genus, and decreased abundance of bacteria of the *Firmicutes* phylum and was seen in AD cases when compared to healthy controls across several of the 11 studies included in this review (6).

### Limitations

There are some notable limitations to our study. Firstly, we included one microbial trait (i.e. *G. Bifidobacterium* [AB]) for which the associated genetic variant did not meet the traditional genome-wide p-value threshold in the GWAS meta-analysis. However, this is the most well characterised genetic variant that is consistently associated with bacteria in the entire *Bifidobacterium* taxonomical tree (31, 32). Secondly, whilst the third MR assumption (i.e. IV3, “no horizontal pleiotropy”) is untestable, we can provide evidence against the likelihood of unbalanced pleiotropy using methods such as the MR-Egger, weighted median and weighted mode methods. As a field, we are currently unable to use such methods appropriately in main analyses given the very small numbers of robustly related genetic variants associated with microbial traits. However, what we have demonstrated here is that the application of MR, coupled with sensitivity analyses such as colocalisation and a more manual approach to investigating horizontal pleiotropy, can go some way to challenging the validity of this MR assumption. As GWAS of the gut microbiome are heterogenous (from the sampling through to the analysis), until larger GWAS of the gut microbiome are conducted in a homogenous way that provides a greater number of more reliable genetic signals, this will remain a substantial limitation in applying MR to understanding the role of the gut microbiome on health outcomes. Third, assessment of cognitive function measures in UK Biobank did not use the gold standard assessments carried out by trained psychologists and instead used fully automated touchscreen assessments. Thus, these measures lack validity and reliability. That said, previous studies have shown they are likely a good indicator of cognition (33). Lastly, these analyses were undertaken with only individuals of European ancestry; therefore, these results may not be generalisable to other populations.

## Conclusions

In conclusion, whilst our analyses initially provided evidence that features of the gut microbiome may influence the risk of AD and cognition, further sensitivity analyses indicated that these results were likely not reflective of causality. Future applications of MR to understand the role played by the gut microbiome on health and disease need to take more caution in appropriately applying and interpreting MR findings and consider combining evidence from MR analyses with findings from other study designs, which have different key sources of bias, to improve causal inference.

## Supporting information

Supplement 2 - STROBE MR checklist

## Data Availability

All the data referred to in the manuscript is available online. Summary GWAS data for both Alzheimer's Disease GWASs was obtained from Jansen et al. (2019) at https://pmc.ncbi.nlm.nih.gov/articles/PMC6836675/. Summary GWAS data for fluid intelligence and reaction time were obtained from IEU GWAS pipeline (https://data.bris.ac.uk/data/dataset/pnoat8cxo0u52p6ynfaekeigi).

## Abbreviations

AD: Alzheimer’s disease
AB: *Bacteria* abundance
P/A: *Bacteria* presence or absence
IV1: Core MR assumption 1
IV2: Core MR assumption 2
IV3: Core MR assumption 3
FGFP: Flemish Gut Flora Project
FoCus: Food-Chain Plus
GWAS: Genome-wide association study
IGAP: International Genomics of Alzheimer’s Project
IVW: Inverse variance weighted
LD: Linkage disequilibrium
MRC: Medical Research Council
MR: Mendelian randomisation
PGC: Psychiatric Genomics Consortium
SNP: Single nucleotide polymorphism
SD: Standard deviation
SE: Standard error
STROBE-MR: Strengthening the Reporting of Observational Studies in Epidemiology MR

## Declarations

### Ethics

All data used for this manuscript were publicly available, where each contributing study had appropriate ethical approval and consent from participants in place.

### Consent for publication

Not applicable.

### Availability of data and materials

Data sharing i s not applicable to this article, as no new datasets were generated for the current study. However, we have provided links to all resources used. Summary GWAS data for both AD GWASs was obtained from Jansen et al. (2019) at https://pmc.ncbi.nlm.nih.gov/articles/PMC6836675/.

Summary GWAS data for fluid intelligence and reaction time was obtained from IEU GWAS pipeline (https://data.bris.ac.uk/data/dataset/pnoat8cxo0u52p6ynfaekeigi). The statistical code for analyses is available on GitHub (https://github.com/genrmonaghan/Alzheimer-s-

Gut-Microbiome-MR.git).

### Contributions

EA and KW were responsible for the conception and design of this work.

GM and CH were responsible for the analysis.

GM, EA, and KW were responsible the interpretation of the data.

GM was responsible for writing the paper which was revised by GM, CH, EA, and KW.

## Acknowledgements

We would like to take this opportunity to thank Dr Panagiota Pagoni for her help with accessing the data and IEU Open GWAS.

