## Supplement 1 - Information for "The causal relationship between gut microbiome composition and Alzheimer’s disease: A two-sample Mendelian randomization analysis"

**Supplementary Information 1**

Reaction time was based on 12 rounds of the card-game 'Snap', where the participant is shown two cards at a time and, if both cards are the same, the participants press a button-box that is on the table in front of them as quickly as possible. Reaction time was measured as the mean time to correctly identify matches in milliseconds (further information is available at https://biobank.ctsu.ox.ac.uk/crystal/field.cgi?id=20023). Fluid intelligence was assessed using a touch screen questionnaire, which measured the participants capacity for problem solving based upon logic and reasoning ability, independent of acquired knowledge. Scores ranged from 0 to 14 based on the number of correctly answered questions in 2 minutes (further information is available at <https://biobank.ctsu.ox.ac.uk/crystal/field.cgi?id=20016>) (3).

**Supplementary Information 2**

The MR-Egger method relaxes the “no horizontal pleiotropy” assumption (i.e. IV3) and allows a non-zero intercept (which would indicate evidence for horizontal pleiotropy). The weighted median method requires only half of the SNPs to be valid instruments (i.e. no confounders of the instrument-outcome association, exhibit no horizontal pleiotropy, and have a robust association with the exposure) for the causal effect estimate to be considered unbiased. The weighted mode method clusters SNPs based on the similarity of their causal effects, weighted by the inverse variance of each SNPs outcome effect, then assumes that the most common causal effect is consistent with the true causal effect and returns the causal effect estimate for the cluster with the largest number of SNPs.
