## Supplement 2 - STROBE MR checklist for "The causal relationship between gut microbiome composition and Alzheimer’s disease: A two-sample Mendelian randomization analysis"

**STROBE-MR checklist of recommended items to address in reports of Mendelian randomization studies**^1^ ^2^

| **Item No.** | **Section** | **Checklist item** | **Page No.** | **Relevant text from manuscript** |
| --- | --- | --- | --- | --- |
| 1 | **TITLE and ABSTRACT** | Indicate Mendelian randomization (MR) as the study’s design in the title and/or the abstract if that is a main purpose of the study | 1 | Title is “The causal relationship between gut microbiome composition and Alzheimer’s disease: A two-sample Mendelian randomization analysis” and the abstract contains details of the purpose and design of the study. |
|  | **INTRODUCTION** |  | 2-3 |  |
| 2 | **Background** | Explain the scientific background and rationale for the reported study. What is the exposure? Is a potential causal relationship between exposure and outcome plausible? Justify why MR is a helpful method to address the study question | 2 | The background and rationale for the study is described in paragraphs 1, 2, 3, and 4 of the introduction. The plausibility of the potential causal relationship between the exposure and outcome is explained in paragraph 2. The justification of why MR is helpful to address the study question is in paragraphs 3 and 4. |
| 3 | **Objectives** | State specific objectives clearly, including pre-specified causal hypotheses (if any). State that MR is a method that, under specific assumptions, intends to estimate causal effects | 2 | The objectives of the study are outlined in paragraph 4 of the introduction |
|  | **METHODS** |  | 3-7 |  |
| 4 | **Study design and data sources** | Present key elements of the study design early in the article. Consider including a table listing sources of data for all phases of the study. For each data source contributing to the analysis, describe the following: | 3-7 | The study design is explained on page 3 in the first paragraph of the methods. The data sources are described across pages 3 and 4. |
|  | a) | Setting: Describe the study design and the underlying population, if possible. Describe the setting, locations, and relevant dates, including periods of recruitment, exposure, follow-up, and data collection, when available. | 3-4 | The study design is explained on page 3 in the first paragraph of the methods. The data sources are described across pages 3 and 4 (which includes an overview of the participants, criteria for inclusion and selection of studies and participants, and the number of cases and controls). |
|  | b) | Participants: Give the eligibility criteria, and the sources and methods of selection of participants. Report the sample size, and whether any power or sample size calculations were carried out prior to the main analysis | 3-4 | The data sources are described across pages 3 and 4 (which includes an overview of the participants, criteria for inclusion and selection of studies and participants, and the number of cases and controls). |
|  | c) | Describe measurement, quality control and selection of genetic variants | 4 | The genetic measurements and the selection of genetic instruments are on page 4 of the methods. |
|  | d) | For each exposure, outcome, and other relevant variables, describe methods of assessment and diagnostic criteria for diseases | 3-4 | The methods of assessment and measurement of both the outcome and exposure are on pages 3 and 4 of the methods. |
|  | e) | Provide details of ethics committee approval and participant informed consent, if relevant | 3-4 | Information on ethical approval is on pages 3 and 4 of the methods. |
| 5 | **Assumptions** | Explicitly state the three core IV assumptions for the main analysis (relevance, independence and exclusion restriction) as well assumptions for any additional or sensitivity analysis | 6 | The core MR assumptions are stated on page 6 of the methods. |
| 6 | **Statistical methods: main analysis** | Describe statistical methods and statistics used | 4-7 | The methods are described between pages 4 to 7 of the methods. |
|  | a) | Describe how quantitative variables were handled in the analyses (i.e., scale, units, model) | 3-4 | The units and measurement of the exposures and outcomes are described on pages 3 and 4 in the methods. |
|  | b) | Describe how genetic variants were handled in the analyses and, if applicable, how their weights were selected |  | NA |
|  | c) | Describe the MR estimator (e.g. two-stage least squares, Wald ratio) and related statistics. Detail the included covariates and, in case of two-sample MR, whether the same covariate set was used for adjustment in the two samples | 4-7 | The methodology for MR including sensitivity analyses are described between pages 4 and 7 in the methods section. |
|  | d) | Explain how missing data were addressed |  | NA |
|  | e) | If applicable, indicate how multiple testing was addressed | 5 | Multiple testing was corrected for using Bonferroni correction, as described on page 5. |
| 7 | **Assessment of assumptions** | Describe any methods or prior knowledge used to assess the assumptions or justify their validity | 7 | The F-statistic and R-squared calculations are provided in Supp Table and described on page 7 in the results section. |
| 8 | **Sensitivity analyses and additional analyses** | Describe any sensitivity analyses or additional analyses performed (e.g. comparison of effect estimates from different approaches, independent replication, bias analytic techniques, validation of instruments, simulations) | 5-7 | The sensitivity analyses are described across pages 5 to 7 in the methods section. |
| 9 | **Software and pre-registration** |  |  |  |
|  | a) | Name statistical software and package(s), including version and settings used | 4,6 | R statistical software was used for the analyses and further details on packages used are on pages 4 and 6 in the methods. |
|  | b) | State whether the study protocol and details were pre-registered (as well as when and where) |  | NA |
|  | **RESULTS** |  | 7-9 |  |
| 10 | **Descriptive data** |  |  |  |
|  | a) | Report the numbers of individuals at each stage of included studies and reasons for exclusion. Consider use of a flow diagram | 3-4 | The number of participants included in each exposure and outcome dataset are provided on pages 3 and 4 of the methods. |
|  | b) | Report summary statistics for phenotypic exposure(s), outcome(s), and other relevant variables (e.g. means, SDs, proportions) | 3-4 | The number of participants included in each exposure and outcome dataset are provided on pages 3 and 4 of the methods. The number of cases and controls, where applicable, are provided in Supp Table 1. |
|  | c) | If the data sources include meta-analyses of previous studies, provide the assessments of heterogeneity across these studies | 4 | The details of the meta-analysis used as one of the outcomes for Alzheimer’s Disease (AD) are provided on page 4 of the methods. |
|  | d) | For two-sample MR:  i.  Provide justification of the similarity of the genetic variant-exposure associations between the exposure and outcome samples  ii.  Provide information on the number of individuals who overlap between the exposure and outcome studies | 3-4 | Information on the study populations can be found on pages 3 and 4 of the methods. |
| 11 | **Main results** |  |  |  |
|  | a) | Report the associations between genetic variant and exposure, and between genetic variant and outcome, preferably on an interpretable scale |  | Provided in Supp Table 1. |
|  | b) | Report MR estimates of the relationship between exposure and outcome, and the measures of uncertainty from the MR analysis, on an interpretable scale, such as odds ratio or relative risk per SD difference |  | Provided in Table 1 and Figure 2. |
|  | c) | If relevant, consider translating estimates of relative risk into absolute risk for a meaningful time period |  | NA |
|  | d) | Consider plots to visualize results (e.g. forest plot, scatterplot of associations between genetic variants and outcome versus between genetic variants and exposure) |  | Provided in Figures 1-3. |
| 12 | **Assessment of assumptions** |  |  |  |
|  | a) | Report the assessment of the validity of the assumptions | 5-9 | Sensitivity analyses to access the validity of the assumptions are provided on pages 5 to 7 of the methods and 8 to 9 of the results. |
|  | b) | Report any additional statistics (e.g., assessments of heterogeneity across genetic variants, such as *I^2^*, Q statistic or E-value) | 5-9 | Sensitivity analyses to access heterogeneity are provided on pages 5 to 7 of the methods and 8 to 9 of the results. |
| 13 | **Sensitivity analyses and additional analyses** |  |  |  |
|  | a) | Report any sensitivity analyses to assess the robustness of the main results to violations of the assumptions | 5-9 | Sensitivity analyses are provided on pages 5 to 7 of the methods and 8 to 9 of the results. |
|  | b) | Report results from other sensitivity analyses or additional analyses |  | NA |
|  | c) | Report any assessment of direction of causal relationship (e.g., bidirectional MR) | 4-9 | Bidirectional MR was carried out, Details of the forwards direction are described on pages 4 to 7 of the methods and 8 and 9 of the results. Details of the reverse direction are described on pages 4 and 7 of the methods and pages 8 and 9 of the results. |
|  | d) | When relevant, report and compare with estimates from non-MR analyses |  | NA |
|  | e) | Consider additional plots to visualize results (e.g., leave-one-out analyses) |  | NA |
|  | **DISCUSSION** |  | 9-11 |  |
| 14 | **Key results** | Summarize key results with reference to study objectives | 9 | Key results are summarised in paragraph 1 of the discussion on page 9. |
| 15 | **Limitations** | Discuss limitations of the study, taking into account the validity of the IV assumptions, other sources of potential bias, and imprecision. Discuss both direction and magnitude of any potential bias and any efforts to address them | 10-11 | Limitations are discussed on pages 10-11 in the discussion. |
| 16 | **Interpretation** |  |  |  |
|  | a) | Meaning: Give a cautious overall interpretation of results in the context of their limitations and in comparison with other studies | 9-10 | The interpretation of results can be seen on pages 9 and 10 and the comparison with other studies can been seen on page 10. |
|  | b) | Mechanism: Discuss underlying biological mechanisms that could drive a potential causal relationship between the investigated exposure and the outcome, and whether the gene-environment equivalence assumption is reasonable. Use causal language carefully, clarifying that IV estimates may provide causal effects only under certain assumptions | 2 | Possible mechanisms are briefly described in paragraph 2 of the introduction on page 2, but given our overall conclusions, these are not elaborated on in the discussion or included in any conclusions. |
|  | c) | Clinical relevance: Discuss whether the results have clinical or public policy relevance, and to what extent they inform effect sizes of possible interventions | 9-10 | The relevance of these results is discussed on pages 9 and 10 of the discussion, but given our overall conclusions, |
| 17 | **Generalizability** | Discuss the generalizability of the study results (a) to other populations, (b) across other exposure periods/timings, and (c) across other levels of exposure | 10-11 | The generalisability of these results is discussed in terms of population samples used in MR analyses on pages 10 and 11 in the discussion |
|  | **OTHER INFORMATION** |  |  |  |
| 18 | **Funding** | Describe sources of funding and the role of funders in the present study and, if applicable, sources of funding for the databases and original study or studies on which the present study is based | 11 | Funding statements and acknowledgements are provided on page 11. |
| 19 | **Data and data sharing** | Provide the data used to perform all analyses or report where and how the data can be accessed, and reference these sources in the article. Provide the statistical code needed to reproduce the results in the article, or report whether the code is publicly accessible and if so, where | 3-4 | Data availability and access are provided on pages 3 and 4 of the methods. |
| 20 | **Conflicts of Interest** | All authors should declare all potential conflicts of interest | 11 | Conflicts of interest for those authors which this is applicable is provided on page 11. |

This checklist is copyrighted by the Equator Network under the Creative Commons Attribution 3.0 Unported (CC BY 3.0) license.

1.	Skrivankova VW, Richmond RC, Woolf BAR, Yarmolinsky J, Davies NM, Swanson SA, et al. Strengthening the Reporting of Observational Studies in Epidemiology using Mendelian Randomization (STROBE-MR) Statement. JAMA. 2021;under review.

2.	Skrivankova VW, Richmond RC, Woolf BAR, Davies NM, Swanson SA, VanderWeele TJ, et al. Strengthening the Reporting of Observational Studies in Epidemiology using Mendelian Randomisation (STROBE-MR): Explanation and Elaboration. BMJ. 2021;375:n2233.
